# The Risk of Bias in Iranian Randomized Controlled Trials Included in Cochrane Reviews

**DOI:** 10.1101/2020.08.02.20166728

**Authors:** Ali Kabir, Ahmad Sofi-Mahmudi, Arman Karimi Behnagh, Vahid Eidkhani, Hamid Reza Baradaran, Payam Kabiri, Ali Akbar Haghdoost, Bita Mesgarpour

## Abstract

**Background:** Randomised controlled trials (RCT) provide the highest level of evidence among interventional studies. However, RCTs may be susceptible to the risk of bias (RoB). Therefore, systematic reviews appraise the RoB in all included studies in a review by using evaluation tools. This study aimed to describe the main characteristics of RCTs conducted in Iran and included in Cochrane Reviews (CRs) by focusing on their RoB.

**Methods:** We searched “Iran” by selecting the “Search All Text” field and “Review” in the *Cochrane Database of systematic Review* within Ovid. We retrieved CRs that included Iranian controlled trials. We selected trials only if they had involved human subjects, described as a controlled clinical trial, included in CRs and therefore the review authors assessed their RoBs. The characteristics of trials have been extracted by looking at the table “Characteristics of included studies” in each CRs as well as the relevant papers. To addressing RoB, we collected the judgment of the review authors according to the table of RoB assessments in CRs.

**Results:** There were 1166 Iranian RCTs included in 571 CRs. From all these studies, 44.9% were at low RoB for random sequence generation, 20.8% for allocation concealment, 32.3% for blinding of participants/personnel, 36.5% for blinding of outcome assessors, 56.3% for incomplete outcome data, 41.3% selective reporting, and 53.8% for other bias.

**Conclusion:** RoB was mainly high or unclear in Iranian RCTs. Special attention must be paid to methodological quality of RCTs in Iran accordingly.

## Introduction

Bias or systematic error can be resulting in over or under-reporting of the treatment effects. Although randomised controlled trials (RCTs) are regarded as the gold standard of clinical research designs, they are frequently at risk of flaws in the design, conduct, analysis, and reporting.^1^ Systematic reviews, especially the ones that focus on intervention studies, mainly include RCTs; however, the quality of RCTs has been a general concern.^2^ Low quality included RCTs in a systematic review is a potential limitation and may result in unreliable effect estimates.^3^ The attempt for quality improvement of trials has been initiated from more than two decades ago, and the Consolidated Standards for Reporting Trials (CONSORT) were developed accordingly.^4^ Nevertheless, lack of adoption the standards by a journal, low adherence to the guideline following its adoption, insufficient training of researchers, poor research governance practices and process may influence the quality of conducting as well as reporting of trials.^5-8^

The quality of RCTs have been frequently studied based on the published journals,^9, 10^ within a specific subject,^11, 12^ in a particular year,^13^ and rarely, in a specific country.^14, 15^ However, these studies have mainly assessed the quality of reporting trials by using CONSORT statement and therefore, rely on the information retrieved from the reports.

Cochrane reviews (CRs) are considered the gold standards of systematic reviews because of implementing the highest standards of quality on conduct and reporting.^16^ CRs are publishing in the Cochrane Database of Systematic Reviews (CDSR), which is the most recognised journal and database of systematic reviews and meta-analyses in healthcare. It is part of the Cochrane Library and includes all Cochrane Reviews (and protocols) prepared by authors who register titles with one of the Cochrane Review Groups.^17^ Each Cochrane Review Group focuses on a specific topic area, which supports Cochrane review authors with methodological and editorial issues. Cochrane recommends a specific tool for assessing the risk of bias (RoB) in each included studies, launched in 2008, updated in 2011 and revised in 2019.^2, 18^

The Iranian Registry of Clinical Trials (IRCT) was established as a WHO primary registry at the end of 2008^19^ and the number of registrations is substantially increased to 25,000 in June 2020. Insufficient quality of reporting Iranian RCTs have been studied before;^20^ however, we have not found any evaluation of the quality of Iranian RCTs in terms of risk of bias assessment. Such evaluation may be useful for many other countries like Iran to understand where they have been stand for the quality of their RCTs.

The purposes of this study were to provide an overview of the characteristics and risk of bias in RCTs conducted in Iran and included in CRs, as well as to identify items where improvement is most required.

## Methods

We searched “Iran” by selecting the “Search All Text” field and “Review” in the *Cochrane Database of Systematic Reviews* within Ovid on September 30, 2019. We screened Cochrane reviews to identify those, which have included Iranian RCTs. We extracted the main characteristics of the eligible CRs, which includes DOI, publication year, Cochrane Review Group, number of Iranian and total trials included, and the total population of included trials. We selected trials only if they had involved human subjects, described as a controlled clinical trial, included in CRs and therefore, Cochrane review authors assessed their RoBs.

Four independent authors (AK, AKB, VE, AS) extracted the characteristics of each trial by looking at the table “Characteristics of included studies” in each CRs and the relevant published paper where data were provided insufficient. We gathered data for two features of trials: 1) the characteristics of a trial’s paper including the first author, the title, the name of the journal, publication year and the language of paper; and 2) the characteristics of trial consisting of the type of intervention, the trial start and end dates, city, province, number of involved centre(s), sample size, type of assignment and the RoB assessments. Randomised controlled trial, quasi-experimental, crossover RCT, cluster RCT, non-randomized and other were considered as RCT assignments.

Cochrane reviews appraise the RoB in all studies in a review by using a specific domain-based evaluation tool. This comprises a judgment for each item involves the RoB as ‘Low risk’, ‘High risk’ or as ‘Unclear risk’ where there is lack of information or uncertainty over the potential for bias. To describe RoB assessments, we extracted the judgment of the review authors for each item according to the RoB assessments table in the CRs. The standard format of Cochrane RoB assessment consists of seven items: random sequence generation, allocation concealment, blinding-performance bias, blinding-detection bias, incomplete outcome data (attrition bias), selective outcome reporting, and other biases. If blinding was reported in other formats like subjective and objective outcomes or assessor, analyst, participants and/or caregivers, we considered an aggregated assessment; if one or more subcategories of blinding item had been reported as high or low risk of bias, we considered the category of blinding as high or low risk of bias. Otherwise, we reported the blinding bias as unclear. Due to diversity of other types of reported bias in each CRs, such as funding bias, intention to treat bias, sample size bias, for-profit bias, and power calculation bias, we aggregated those as other biases where appropriate.

We examined the trend of RoB for Iranian RCTs to explore the role of time in improvement of methodology and reporting. We have not assessed the overall risk-of-bias judgement for the result since it is not favourable in this tool and has not recommended.

### Statistical analysis

We analysed extracted data using R v3.6.0 (2019-04-26) (R Foundation for Statistical Computing, Vienna, Austria. http://www.R-project.org/). We reported frequencies and percentages to present the characteristics of RCTs.

## Results

Among 571 retrieved CRs, 1894 Iranian RCTs were located, 1166 of them had been included in the relevant CRs, and their RoB had been assessed. The reasons for exclusions of Iranian RCTs in the retrieved CRs were mostly due to awaiting for assessment (20.6%) and to be on-going trials (20.6%) (Figure 1).

**Figure 1.**
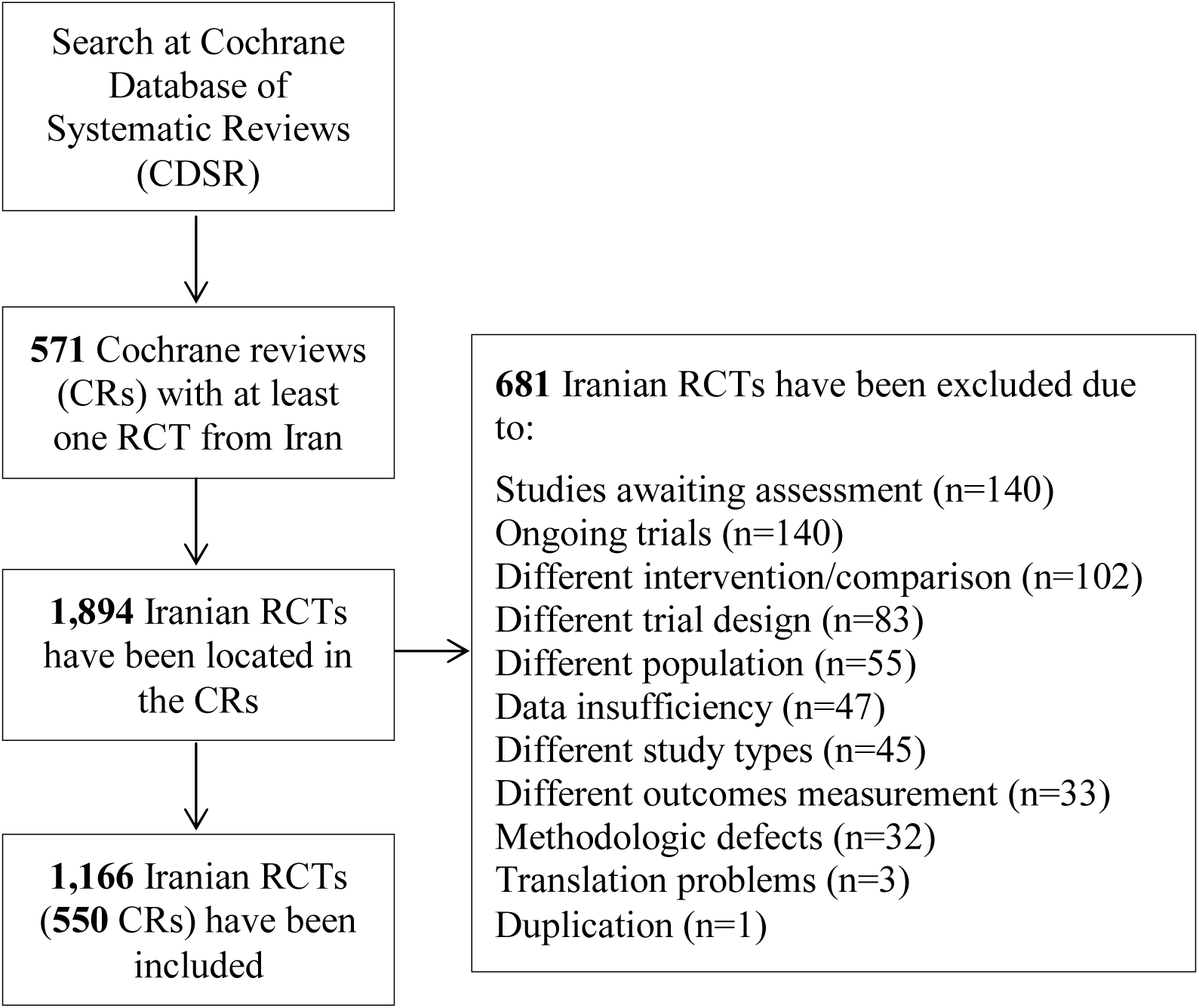
flow diagram of search for Iranian RCTs in Cochrane Reviews.

CRs were included 15,894 RCTs (with 5,461,452 population) in total, which only 7.3 % of those (2.7% of the population) were conducted in Iran. However, the population of five CRs was not reported and is not included in the analysis. CRs were published from 1970 to 2018, which 63.7% of them were published after 2013. Those CRs were managed by 50 Cochrane Review Groups (Figure 2). The majority of Iranian RCTs were in the fields of Pregnancy and Childbirth, Gynaecology and Fertility, and Skin and Oral Health with 250 (21.4%), 158 (13.5%) and 70 (6.0%) studies, respectively. A Cochrane Review published in 2017 by a group of authors from Madrid in the Cochrane Skin Group, entitled “Interventions for Old World cutaneous leishmaniosis” with 27 out of 49 included RCTs from Iran had the highest number of Iranian trials (Heras-Mosteiro, 2017). There were no reviews in the Cochrane HIV/AIDS, Lung Cancer, Methodology Review, STI and Urology groups, which included Iranian RCTs. Two RCTs were included in two Cochrane reviews and RoB assessments in one of those were not consistent.

**Figure 2.**
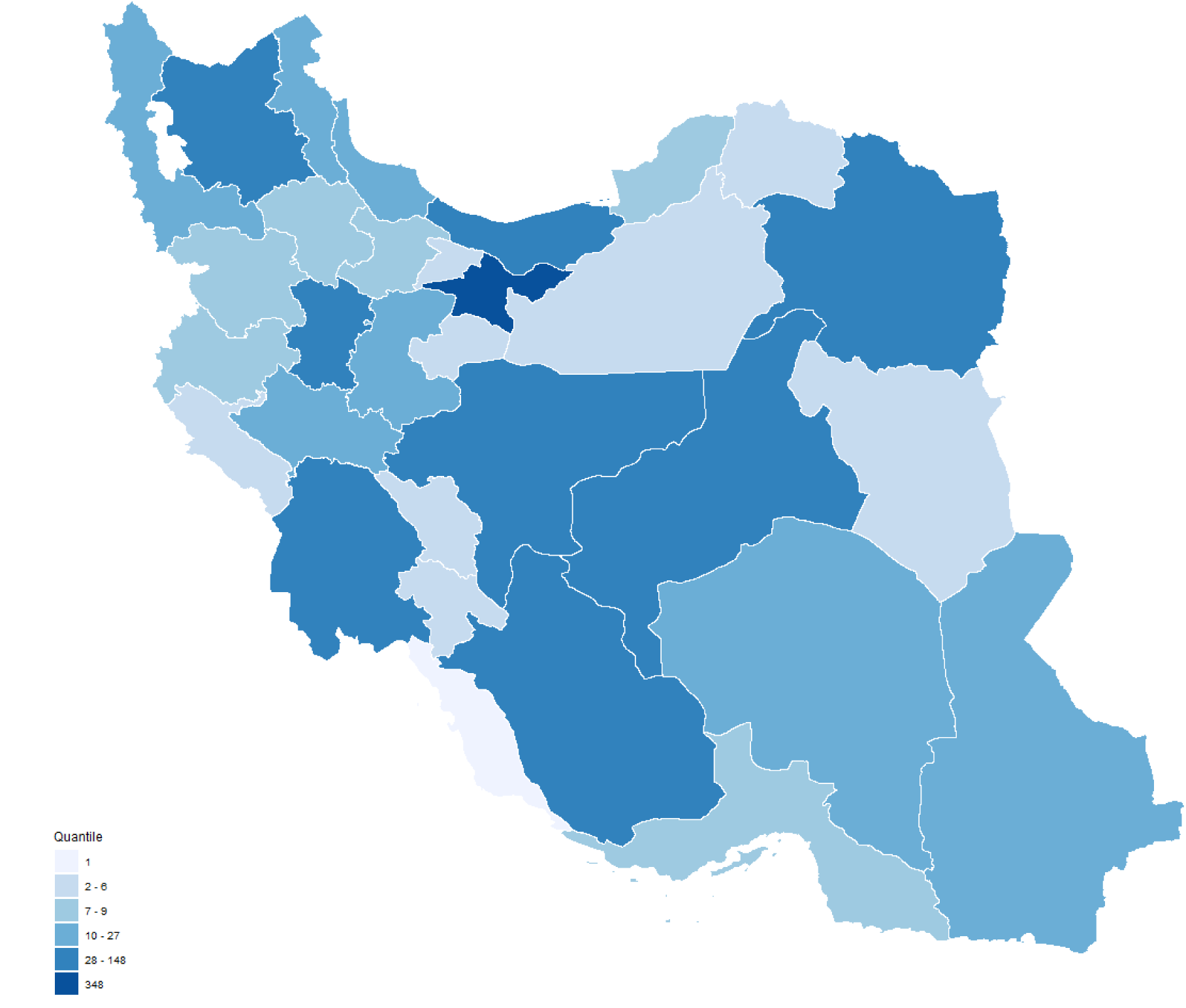
Distribution of conducting sites of Iranian RCTs.

These RCTs were mainly published between 2003 and 2016 (81.0%) in 60 cities. The distribution of the location of the conducting centres was not homogenous, and Tehran, Isfahan, and Fars provinces had the highest number of RCTs with 348 (29.8%), 148 (12.7%), and 97 (8.3%) studies, respectively (Figure 3). The included RCTs appeared in 548 peer-reviewed journals, 8 Conference Proceeding and 4 in Iranian Registry of Clinical Trials (IRCT) with notable distribution in local journals (…%) and in English (90.1%). The highest numbers of publications were in Journal of Research in Medical Sciences, Isfahan University of Medical Sciences (29), International Journal of Gynecology & Obstetrics (29) and Iranian Red Crescent Medical Journal (28).

**Figure 3.**
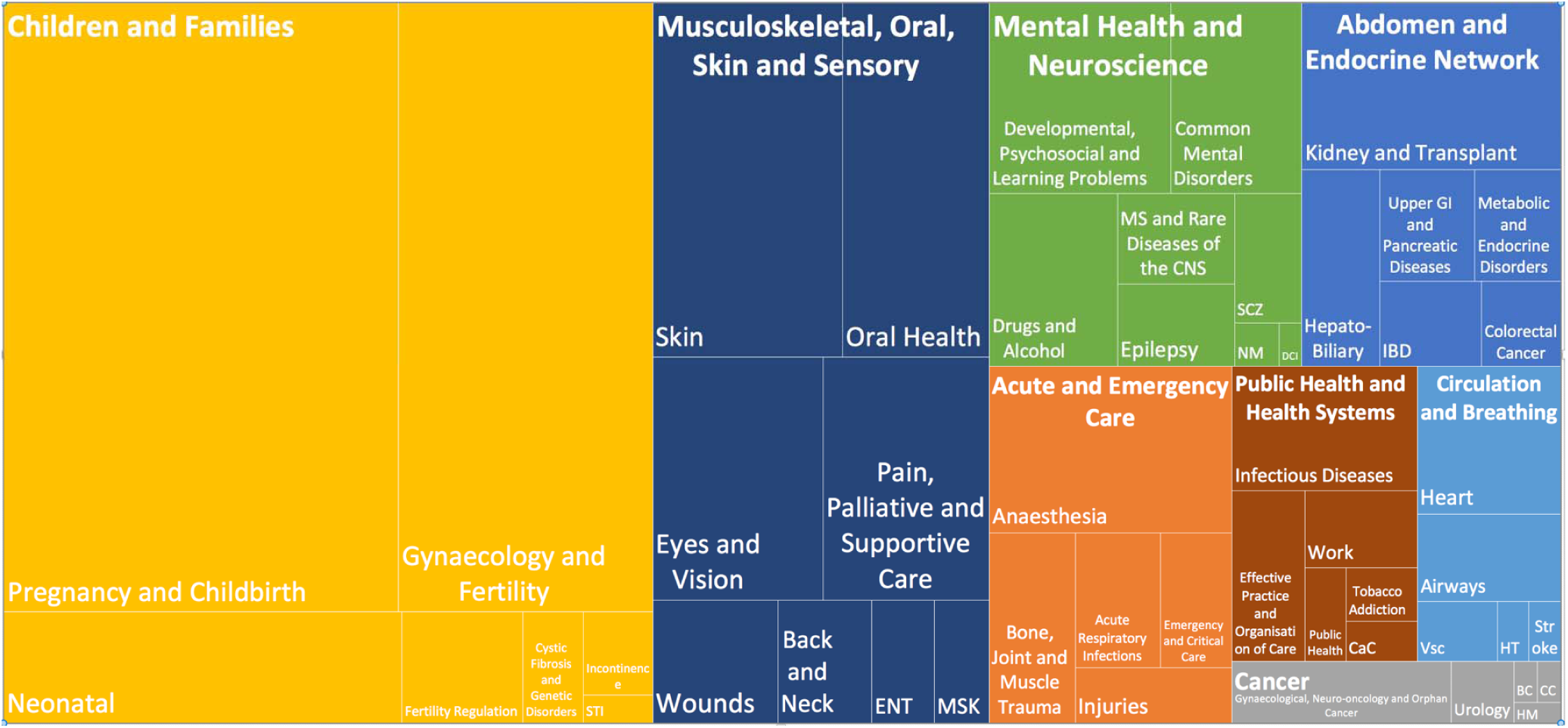
Distribution of Iranian RCTs in the Cochrane reviews based on Cochrane Review Groups. BC: Breast Cancer; CaC: Consumers and Communication; CC: Childhood Cancer; DCI: Dementia and Cognitive Improvement; HM: Haematological Malignancies; HT: Hypertension; MSK: musculoskeletal; NM: Neuromuscular; SCZ: Schizophrenia; Vsc: Vascular

Assignments were randomised controlled clinical trial (RCT) in 1040 (89.2%), and quasi-experimental in 26 (2.2%) studies. The assignments were unclear in one study. The most common targets were treatment (47.3%) and supportive care (24.2%).

The majority of studies (83.6%) had a sample size of between 30 and 200. The median of sample size was 80, and four studies had a sample size over than 1,000. However, 25.5% of studies had equal to or less than 50 participants in their trial. The highest and the lowest number of sample sizes were 12514 and 9, included in Airways and Public Health Review Groups, respectively. We could not find the full text of one article. Table 1 represents the frequency of sample sizes. From a methodological point of view, 1077 (92.4%) studies reported that they had at least one arm as the control group which 299 (25.6%) of them this arm received placebo. About blinding, 414 (35.5%) studies were labelled as double-blinded, 28 (2.4%) studies labelled as triple-blinded, and 266 (22.8%) studies considered no method for blinding in their design.

**Table 1.** Frequency of each sample size group in Iranian RCTs.

| Sample size group | N (%) | Cumulative |
| --- | --- | --- |
| 9-20 | 34 (2.9) | 34 (2.9) |
| 21-40 | 160 (13.7) | 194 (16.6) |
| 41-60 | 232 (19.9) | 426 (36.5) |
| 61-80 | 176 (15.1) | 602 (51.6) |
| 81-100 | 169 (14.5) | 771 (66.1) |
| 101-150 | 188 (16.1) | 959 (82.2) |
| 151-200 | 87 (7.4) | 1046 (89.6) |
| 201-300 | 59 (5.1) | 1105 (94.7) |
| 301-400 | 29 (2.5) | 1134 (97.2) |
| 401-500 | 9 (0.9) | 1149 (98.1) |
| 501-1000 | 18 (1.5) | 1161 (99.6) |
| 1001-12514 | 4 (0.4) | 1165 (100) |

Not all domains of RoB tool were assessed in all reviews. For example, Cochrane reviewers had assessed random sequence generation and allocation concealment in 1134 and 1122 out of 1166 RCTs, respectively. “Blinding (performance bias)” domain had the highest rate of high-risk of bias (22.9%) and “Incomplete outcome data” had the highest rate of low-risk of bias (56.3%). The judgement of at least one risk of bias domain as “unclear” was found in 87.3% of included randomised clinical trials (931/1066). Overall assessment of included RCTs for each domain of RoB tool are illustrated in Figure 4. However, random sequence generation and incomplete outcome data domains showed a continuous improvement regarding low RoB during 2002-2017 (Figure 5, The analysis for all domains has been provided in Appendix 1).

**Figure 4.**
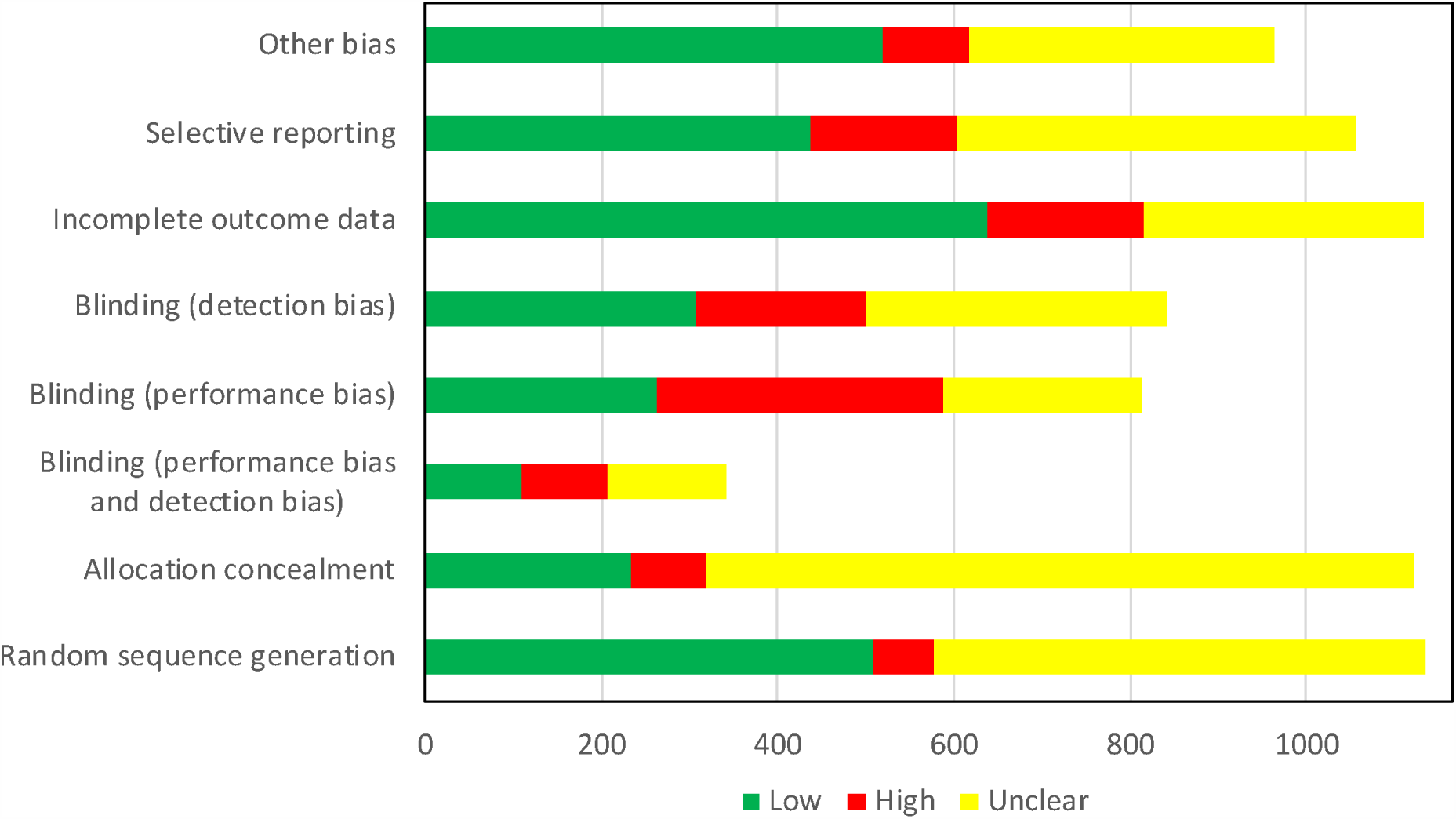
Overall Risk of Bias assessment of Iranian RCTs included and evaluated in Cochrane Reviews.

**Figure 5.**
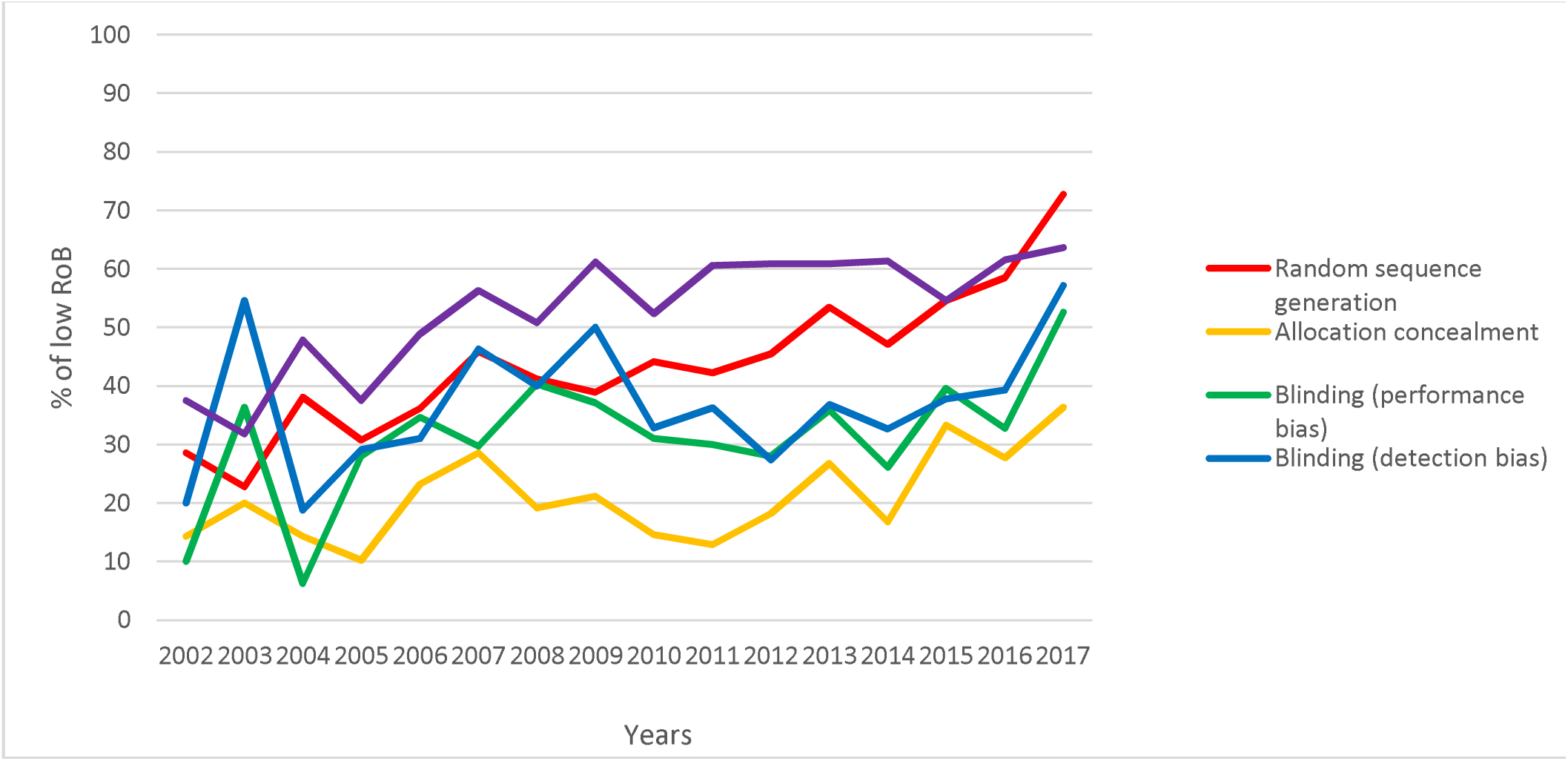
The trend of low RoB for four domains of Cochrane tool in the Iranian RCTs during 2002-2017.

## Discussion

In this study, we tried to address the quality of the Iranian RCTs included in Cochrane systematic reviews based on the Cochrane reviewers’ evaluations. To the best of our knowledge, this is the first study using Cochrane RoB tool to evaluate the quality of RCTs in a national scope. We found out that in the majority of the RoB domains, mostly the ones associated with study design, including random sequence generation, allocation concealment, and blinding, Iranian RCTs have poor quality and mostly assessed as at high or unclear RoB.

There are several studies, which evaluate the quality of Iranian RCTs based on the CONSORT checklist. Although these studies basically tried to consider RCTs of particular subjects or fields but the findings were consistent and indicative of the poor methodological quality and reporting of the Iranian RCTs from a methodological point of view.^21-24^

In this study, we demonstrated that 44.9% of the Iranian RCTs were evaluated as low-risk of bias in random sequence generation and the rest of studies were not be able to provide sufficient data for showing if they could possibly perform a correct form of randomisation. Likewise, in other studies, it was reported that many Iranian RCTs failed to report their method of randomisation in their studies (from 35.5% to 98.7%) so their randomisations were considered to be annulled.^21, 25-27^ This issue is also evident in RCTs conducted in other countries. For example, a study on Saudi Arabian RCTs showed similar results to our study regarding randomisation (44% at low risk of bias).^15^ In a study on 1286 RCTs included in CRs, less than half of them were at low risk of bias.^28^ It means this issue is a global issue and not specific to Iran so that a global effort is needed to enhance RCTs quality in this domain.

Blinding was the other domain divided into two subdomains: blinding of performance and blinding of detection. The first one tries to consider the blinding of the participants and the latter one represents the blinding of the outcome assessor(s) and the analyst(s). Regarding blinding of performance, only 262 (32.3%) included studies were assessed as low-risk while in 299 studies placebos were used as a method for blinding of participants and the caregivers. This distance between the low-risk assessed studies and the number of studies using placebo shows that Iranian researchers might fail to comprehend the mechanism of implementing placebo as an instrument to hide intervention from the participants or, at least, they were not able to describe the mechanism in a proper way; therefore, Cochrane reviewers were not persuaded with the explanation. Moreover, in the other subdomain of blinding, detection, similar to the other subdomain but slightly higher, one-third of studies catches the low-risk evaluation mark. One explanation can be the Iranian people’s low tendency to participating in fully blinded researches. The other reason can be due to failure in reporting details of the study, and therefore, this put the reviewers into trouble of assessing bias. Other studies which evaluated Iranian or RCTs from other countries have also pointed out that blinding is usually not adequately performed.^15, 28, 29^

The next domain is attrition bias, which overall has 56.3% of all assessed as low-risk of bias. It seems that the number of patients losing to follow up is low in the Iranian RCTs. Like other studies assessing Iranian RCTs, here in this domain, Iranian researchers were (to some extent) successful in providing data on the patients leaving the study during the active phase. However, international studies show that many RCTs are at low-risk in this domain.^15, 28^ The other domain was selective outcome reporting which the number of low-risk and unclear-risk assessed studies were close, 436 (41.3%) and 453 (42.9%), respectively. One reason to answer why the number of unclear-risk assessed in high is that most of the Iranian authors had not registered their studies in organisations like ClinicalTrials.gov or IRCT. Therefore, there were not any protocols of the studies before the final results would be published. So, the reviewers might be in trouble to evaluate this domain based on the outcomes reported in the final article. Generally, selective outcome reporting and other bias domains are hard to assess; ^30, 31^ therefore, these results should be interpreted with caution.

We found out that most of the Iranian RCTs suffer from the low sample size. The majority of studies had a sample size of less than 60. Furthermore, the distribution of the conducting sites is far heterogeneous, and many studies are single-centre RCTs conducting in Tehran (the capital of Iran). However, this is not surprising as Tehran has many more universities compared to other cities and provinces and leading medical universities are based in Tehran. This highlights a need for encouraging to conduct more collaborative and multi-centre high-quality studies through making good research policies.

As Chalmers and Glasziou have pointed out, there is a considerable amount of research waste in medical research, estimating to be a total of 85%.^32^ This waste could occur in many aspects and phases, namely relevancy of the research question to the patients and the physicians, appropriateness of the study design, accessibility of full publication, and unbiased and usable reporting. Based on our results, Iranian RCTs suffer from all these four issues. Some of these, such as low-quality reporting, could be easily avoided. With the introduction of CONSORT statement in 1996,^4^ we can demonstrate a higher percentage of studies with low-risk of bias in randomisation, blinding of outcome assessors, incomplete data reporting, and other bias. However, due to the high rate of unclear risk of bias in every domain, it can be an alarming sign for low-quality of reporting of the Iranian RCTs.

One advantage of this study is that the studies were evaluated by third-party reviewers who did not possess any tendency to evaluate the studies in a particular direction (bias). However, there was discrepancy in risk of bias (RoB) judgments across Cochrane reviews for one study appeared in two different Cochrane review. This issue has been raised in the previous assessments of RoB based on Cochrane tool as well.^33^

## Conclusion

In this study, we showed that the Iranian researchers have a long way ahead in order to conduct a well-designed RCT and report it. Although a great deal of improvement was observed from initial RCTs to the most recent studies, however in some domains like allocation concealment and blinding special attention must be paid to methodological training.

## Data Availability

Data will be available upon request.

